# Sex-specific differences in resting-state functional brain activity in pediatric concussion

**DOI:** 10.1101/2021.07.14.21260531

**Authors:** Bhanu Sharma, Carol DeMatteo, Michael D. Noseworthy, Brian W. Timmons

## Abstract

**Importance:** Pediatric concussion has a rising incidence and can lead to long-term symptoms in nearly 30% of children. Resting state functional magnetic resonance imaging (rs-fMRI) disturbances are a common pathological feature of concussion, though no studies have examined sex-differences with respect to this outcome. Despite known sex-differences in how pediatric concussion presents, females have remained understudied in rs-fMRI studies, precluding a sex-specific understanding of the functional neuropathology of pediatric concussion.

**Objective:** To provide the first insights into sex-specific rs-fMRI differences in pediatric concussion.

**Design, setting, and participants:** Secondary data analysis of rs-fMRI data collected on children with concussion recruited from in a pediatric hospital setting, with control data accessed from the open-source ABIDE-II database. In total, 27 children with concussion (14 females) approximately one-month post-injury and 1:1 age- and sex-matched healthy controls comprised our sample.

**Exposure:** Patients received a physician diagnosis of concussion. ABIDE-II healthy controls were typically developing.

**Main outcomes & measures:** Seed-based (which permitted an examination of whole-brain connectivity, fitting with the exploratory nature of the present study) and region of interest (ROI) analyses were used to examine sex-based rs-fMRI differences. Threshold-free cluster enhancement (TFCE) and a family-wise error (FWE) corrected p-values were used to identify significantly different clusters.

**Results:** In comparing females with concussion to healthy females, seed-based analyses (in order of largest effect) showed hypo-connectivity between the anterior cingulate cortex of the salience network and the precuneus (TFCE=1173.6, p=FWE=0.002) and cingulate gyrus (TFCE=1039.7, p-FWE=0.008), and the posterior cingulate cortex (PCC) of the default mode network and the paracingulate gyrus (TFCE=870.1, p-FWE=0.015) and sub-callosal cortex (TFCE=795.4, p-FWE=0.037); hyper-connectivity was observed between the lateral pre-frontal cortex and inferior frontal gyrus (TFCE=1215.4, p-FWE=0.002) and lateral occipital cortex (TFCE=854.9, p-FWE=0.020) and between the PCC and cerebellum (TFCE=791.0, p-FWE=0.038). ROI analyses showed primarily patterns of hyper-connectivity in females. No differences were observed between males with concussion and healthy males on seed-based or ROI analyses.

**Conclusions and relevance:** There are alterations in rs-fMRI in females with concussion at one-month post-injury that are not present in males, which provides further evidence that recovery timelines in pediatric concussion may differ by sex.

**KEY FINDINGS:** *Question:* Are there sex-differences in resting state functional brain activity in pediatric concussion?

*Findings:* Females with concussion show both hyper- and hypo-connectivity between multiple brain regions when compared to healthy age- and sex-matched controls. The same analyses revealed no differences in resting state brain activity when comparing males with concussion to their age- and sex-matched healthy peers.

*Meaning:* There are sex-differences in resting state brain activity in pediatric concussion. This suggests that the functional neuropathology of the injury differs between males and females, which may account for sex-differences in the clinical presentation of pediatric concussion.

## INTRODUCTION

Concussion is a mild form of traumatic brain injury (TBI) that results in altered neurological function after biomechanical impact^1^. In pediatric populations, concussion is of particular concern given that it is one of the most common injuries among children and adolescents^2-4^ and has a rapidly rising incidence in those aged 10-19^5,6^. While the injury is transient for the majority of pediatric patients, between 14-29% experience persistent concussion symptoms^7-9^ (PCS; previously referred to as post-concussion syndrome, marked by symptoms which last in excess of four weeks^1^). PCS can include somatic, cognitive, emotional, and sleep-related features that negatively impact academic outcomes^10^ and health-related quality of life^11,12^.

Brain function in pediatric concussion has been studied to understand the nature and extent of its impairment post-injury, as well as its potential etiological role with respect to concussion symptoms. Studies have measured brain function using resting-state functional magnetic resonance imaging (rs-fMRI), which maps regions of brain activity (by proxy of the blood-oxygen-level-dependent, or BOLD, response) and the relative associations between them in a task-independent manner. Pediatric rs-fMRI studies have shown increased functional connectivity in comparison to healthy controls in widely-studied brain networks within the first-week of injury^13-15^. At one-month post-injury (the expected time of recovery^1^), results of rs-fMRI studies are mixed^13,14,16-18^. With respect to studies of children diagnosed with PCS, one study found that within-network functional connectivity across seven validated brain networks did not differ between children with PCS (n=110) vs. healthy peers (n=20), although select PCS symptoms, sleep impairment, and poorer cognition were associated with connectivity in the concussed cohort^19,20^.

A notable limitation common to many pediatric concussion rs-fMRI studies are the imbalanced samples with respect to sex. Some studies involved male only cohorts^14,16,21^, whereas others had less than 25% female representation in their samples^13,17^; in some cases, data on sex were not reported^18,22^. Only a few studies had samples that approached balance (40-45% female) with respect to sex distribution^15,19,20,23^, though these studies did not stratify their results by sex, instead providing group-level data comparing mixed-sex cohorts of children with concussion to their healthy peers. The most direct data on sex-specific rs-fMRI differences come from a recent study involving adults with PCS^24^. The lack of a sex-specific understanding of rs-fMRI differences in pediatric concussion is a considerable knowledge gap, given that sex, as a biological variable, has been recognized as an understudied yet important consideration in neuroscience^25-27^. Further, a growing body of research demonstrates that concussion presents differently in boys vs. girls^28,29^. For example, a recent cohort study (n=986) found that female adolescents with concussion endorse more symptoms on the 22-item and widely used SCAT5^30^ than concussed males, and are more likely to have a higher total symptom score^31^. Two large-scale, multi-center cohort studies have shown that females have a protracted recovery in comparison to males^28,29^, which align with other clinical data on disparate sex effects in concussion summarized in two recent systematic reviews^32,33^. Therefore, we studied sex-specific rs-fMRI differences in pediatric concussion to address an important knowledge gap, and advance our understanding of how the functional neuropathology of concussion differs between males and females.

## METHODS

### Design

The present study is a secondary analysis of data collected as part of two cohort studies (sharing recruitment methods, inclusion/exclusion criteria, and imaging parameters, as detailed below) on pediatric concussion. Control data were obtained from an open-source pediatric neuroimaging database (detailed below). This study was approved by the Hamilton Integrated Research Ethics Board (https://hireb.ca).

### Participants

Children (aged 9-17) experiencing concussion symptoms were recruited by the clinical study team from sites at or affiliated with McMaster University, including the McMaster Children’s Hospital and associated rehabilitation and sports medicine clinics, as well as through direct referral from community physicians. Children diagnosed with a concussion, and their families, were recruited for an intake assessment. Neuroimaging data were then collected as soon after recruitment as scheduling permitted. For the present and larger studies, exclusion criteria included: i) more severe forms of head injury that required surgery, resuscitation, or admission to the critical care unit, ii) complex injuries involving multiple organ systems, and iii) diagnosed neurological disorder or developmental delay.

Imaging data on healthy children were acquired from the multi-site, internationally compiled, open-source Autism Brain Imaging Data Exchange II (ABIDE-II) database^34^. The ABIDE-II database is comprised of over one-thousand anonymized brains (including 557 healthy controls) collected from 19 sites, primarily in North America and continental Europe, yielding nearly 75 publications to date. Both anatomical and functional scans from the ABIDE-II database were pulled to serve as 1:1 age- and sex-matched typically developing controls for our participants with concussion.

### MRI procedures data acquisition

All children with concussion were scanned at a single site (Imaging Research Centre [IRC] at St. Joseph’s Hospital) using a 3-Tesla GE Discovery MR750 MRI scanner and a 32-channel phased array head receiver coil. Upon entering the IRC, participants (as well as their parents and/or guardians if aged 16 years or younger) were led through an intake questionnaire by the MRI technologist, which informed participants about what to expect during the MRI and to ensure that there were no contraindications to MRI. The MRI technologist situated the participant in the MRI, using foam cushioning to minimize discomfort and motion during the scan, and earplugs were provided. The MRI technologist remained in verbal contact with the participant via intercom throughout the scan.

With respect to MRI data collection, first, a 3-plane localizer with calibration sequences was acquired. Anatomical images were then collected using a 3D inversion recovery-prepped fast SPGR T1-weighted sequence (TR/TE=11.36/4.25ms, flip angle=12°, 512×256 matrix interpolated to 512×512, 22cm axial FOV, 1mm thick). Resting state fMRI involved BOLD imaging (gradient echo EPI, TR/TE=2000/35ms, flip angle=90°, 64×64 matrix, 180 time points, 3mm thick, 22 cm FOV), wherein participants were asked to remain awake, keep their eyes open, and not to think of anything in particular. A B_0_ map was acquired for resting state scans, using the same geometric prescription and a B_o_ mapping tool available on the GE scanner which provides a parametric map of field homogeneity in Hz. In regards to the scanning sequence, the rs-fMRI data were acquired within 10-minutes of entering the MRI, as to avoid motion onset by restlessness later in scans as we have observed in this population. Additional data were collected (including DTI^35^ and task-based fMRI data^36^) as part of the imaging battery, but are not relevant to the present study.

With respect to control data from the ABIDE-II database, only scans with a minimum of 180 time-points were used as age- and sex-matched controls. B_0_ data were not available for healthy controls.

### MRI pre-processing and analyses

Pre-processing of imaging data was performed in CONN 19c^37^ (which draws on some the functionality of SPM12^38^), run on MATLAB R2020a. For concussion data only, given that B_0_ maps were not available for controls, unwarping of functional data was performed outside of CONN using the *epiunwarp* script^39^. Unwarped images were then inputted into the pre-processing pipeline which involved the following steps: 1) Functional realignment, wherein all scans were co-registered to the first image acquired^40^. 2) Slice-timing correction to correct for the inherent sequential process of image acquisition, wherein the functional data is time-shifted and resampled (per sinc-interpolation) to match the time in the mid-point of each TR^41^. 3) Functional data outlier detection using SPM’s Artifact Detection Tool (ART)^38^, wherein outliers are detected based on subject motion (>0.9mm framewise displacement) and relative BOLD signal fluctuations (scans ≥5 standard deviations identified as potential outliers). 4) Direct segmentation and normalization/registration of functional data to MNI space (1mm and 2mm isotropic voxels for anatomical and functional data, respectively), based on posterior tissue probability maps. And 5) spatial smoothing of functional data with a Gaussian kernel of full-width at half-maximum of 6mm. All data were inspected visually after pre-processing, as well as by running CONN’s quality assurance checks.

Next, de-noising procedures were performed in CONN. First, ordinary least squares regression was used to project out noise components (associated with cerebral white matter and cerebrospinal regions^42^, outlier scans^43^, and subject motion^44^) using CONN’s anatomical component-based noise correction procedure (aCompCorr). Subsequently, temporal filtering was performed, filtering out frequencies below 0.008Hz and above 0.01Hz. Data were again inspected visually and per the quality assurance metrics offered by CONN.

Seed-based connectivity and ROI-to-ROI based connectivity measures were then computed for each individual subject; groupwise comparisons (comparing all children with concussion to all control participants, healthy girls to girls with concussion, and healthy boys to boys with concussion) were then performed. Seed regions from four, large-scale, validated and clinically-salient (in pediatric concussion and otherwise) resting-state brain networks were used^45-48^. These included the DMN (seeded at the posterior cingulate cortex [1, -61, 38]), salience network (SN, seeded at the anterior cingulate cortex [0, 22, 35]), fronto-parietal network (FPN, seeded at the lateral pre-frontal cortices), and sensorimotor network (SMN, seeded superiorly at the pre-central gyrus [0, -31, 67]). Given that this study is the first to look at sex-differences in rs-fMRI in pediatric concussion, seed-based analyses were employed to explore the relation between these seeds and all other voxels of the brain; an accompanying ROI-to-ROI analysis was also performed (which examines the associations between 164 regions defined by the Harvard-Oxford atlas).

Cluster-level inferences were made per Threshold Free Cluster Enhancement (TFCE)^49^, which avoids the use of an *a priori* cluster-forming height threshold. More specifically, TFCE scores are weighted based cluster extend (or how broad the cluster is, spatially speaking) and the cluster-height (without examining only at the “peaks” of clusters that survive the threshold that is otherwise arbitrarily set). For each groupwise contrast (as specified above) permutation tests (involving 10,000 permutations) were used to derive a null distribution that the observed effects were then compared to, and a TFCE score associated with family-wise error (FWE) corrected p-value for each cluster was obtained. TFCE has the advantage of being associated with a lower false-positive rate than traditional cluster-size tests based on random field theory^50^.

Between-group contrasts (i.e., all healthy vs. all concussed, healthy males vs. males with concussion, healthy females vs. females with concussion) were set up in CONN 19c as independent samples t-tests. For each contrast and for each seed-region, significantly different clusters were identified using TFCE. The effect sizes associated with each significant cluster were computed, along with a t-score to statistically compare effect size differences at the cluster level between groups.

## RESULTS

Demographic and injury data of the 27 children with concussion and 27 controls are summarized in **Table 1**. Age did not significantly differ between males and females in either cohort, and males and females with concussion had similar PCSS scores (47.8 vs. 41.6, p=0.511) at time of imaging per an independent samples t-test. Patients with concussion were, on average, approximately one-month post-injury (28.8 ± 14.5 days) at time of imaging, and had no history of anxiety, depression, sleep disorder, or psychiatric diagnosis.

**Table 1.**
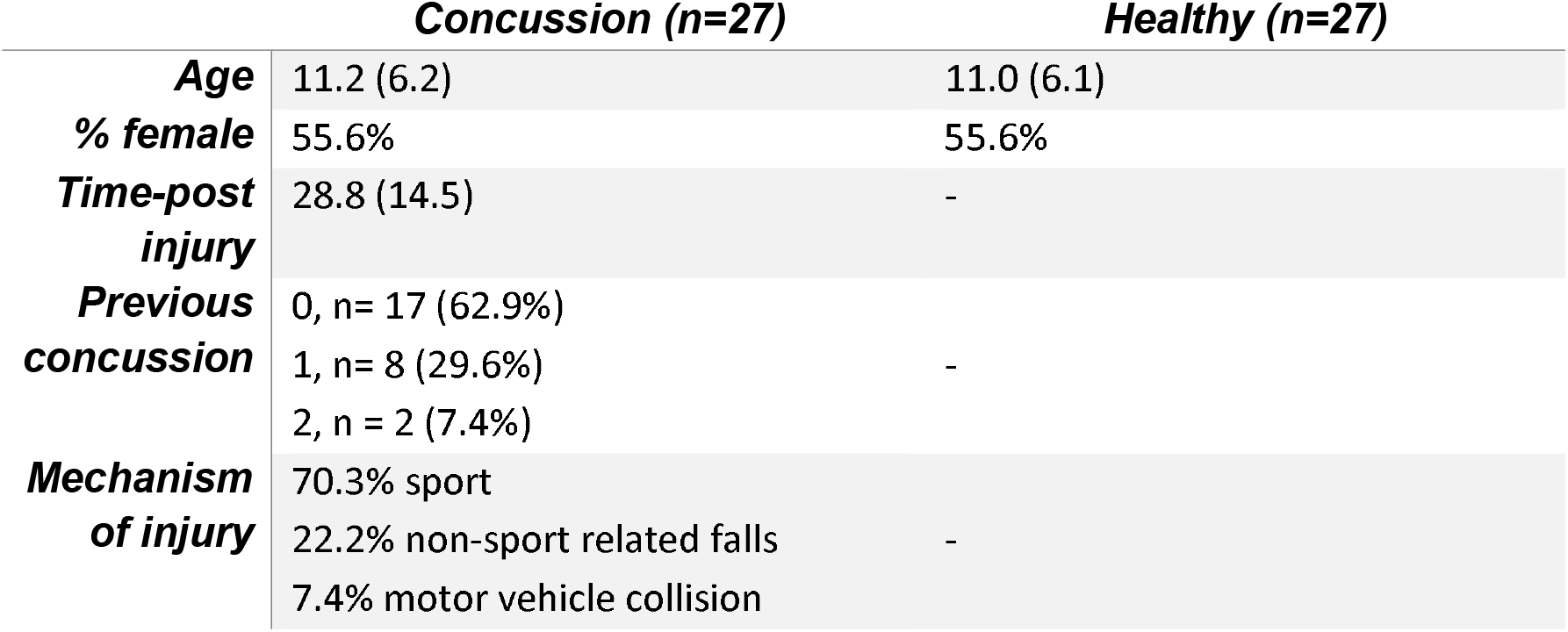
Demographic and injury-related variables.

|  | ***Concussion (n=27)*** | ***Healthy (n=27)*** |
| --- | --- | --- |
| ***Age*** | 11.2 (6.2) | 11.0 (6.1) |
| ***% female*** | 55.6% | 55.6% |
| ***Time-post injury*** | 28.8 (14.5) | - |
| ***Previous concussion*** | 0, n= 17 (62.9%)<br>1, n= 8 (29.6%)<br>2, n = 2 (7.4%) | - |
| ***Mechanism of injury*** | 70.3% sport<br>22.2% non-sport related falls<br>7.4% motor vehicle collision | - |

### Concussion vs. control group comparison

In the concussion cohort, there was significantly reduced functional connectivity (all p-FWE <0.05, with corresponding TFCE scores in **Figure 1**) between the seed-region of the: i) DMN and the superior frontal gyrus (bilaterally), paracingulate gyrus (right), and sub-callosal cortex; ii) SMN and superior frontal gyrus (left), lateral occipital cortex (left), and cuneal cortex (left); iii) SA and precuneous cortex, cingulate gyrus (left) and hippocampus (left), and intra-calcarine cortex (left). Further, there was increased functional connectivity between the seed-region of the FPN and the inferior frontal gyrus (left) and lateral occipital cortex (left); there was also increased functional connectivity observed between the DMN seed and cerebellum (left). These data are depicted in **Figures 1** and **eFigure1**.

**Figure 1.**
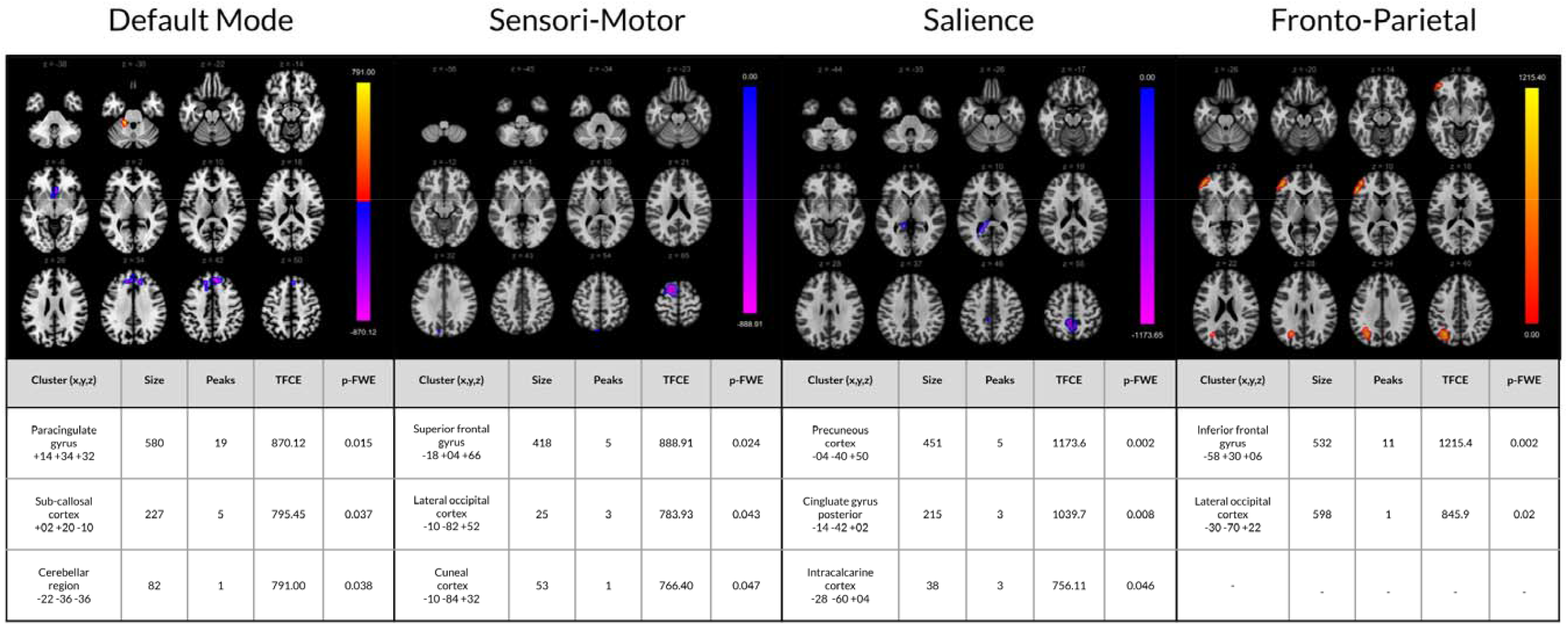
rs-fMRI differences between children with concussion and their healthy peers (mixed-sex cohorts). Clusters (x, y, z) denote standard MNI coordinates at the center of cluster mass, and size represents number of voxels. Only statistics for clusters that survived a p-FWE <0.05 per TFCE are displayed.

At the ROI-to-ROI level, there was a broad pattern of increased functional connectivity between multiple brain regions bilaterally, and fewer instances of decreased connectivity between pairs of ROIs. The pairs of ROIs with significantly reduced connectivity included: the lateral parietal node (left) and supramarginal gyrus (left), superior frontal gyrus (left) and thalamus (right), inferior frontal gyrus (right) and thalamus (left) (see **Figure 2**).

**Figure 2.**
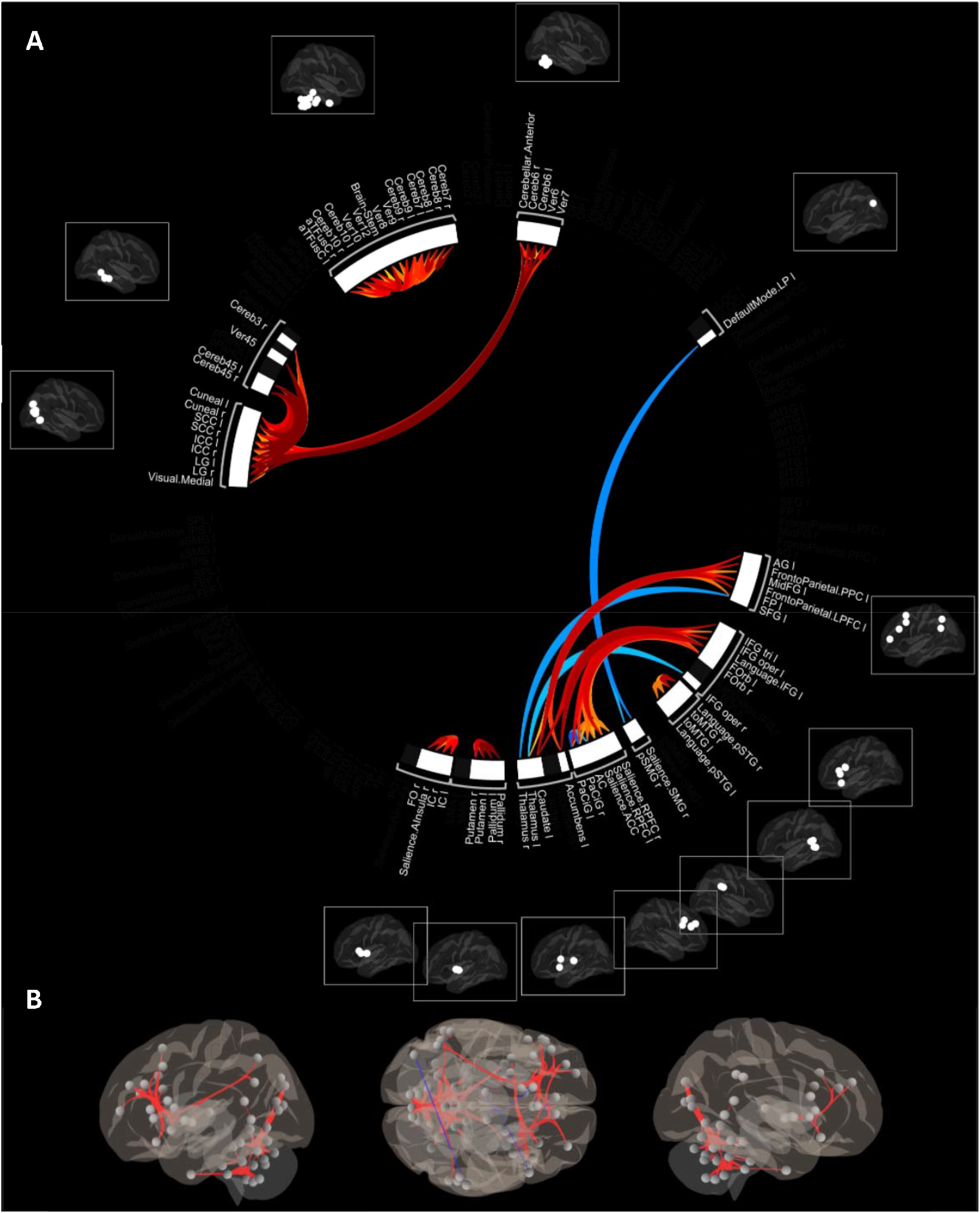
(A) Significantly increased (warm colours) and decreased (cool colours) ROI-to-ROI connectivity in children with concussion in comparison to controls (mixed-sex sample). (B) Depiction of the ROI-to-ROI changes summarized in panel A on a 3D glass brain to more clearly depict altered patterns of connectivity in an anatomically relevant space.

### Healthy males vs. males with concussion

Per both seed-based and ROI-to-ROI level analyses, there were no significant groupwise differences observed between healthy males and males with concussion.

### Healthy females vs. females with concussion

In females, with respect to the DMN, there was increased connectivity between the DMN seed and parts of the cuneal cortex (right), and reduced connectivity between said seed and primarily the insular cortex (left), frontal oribital cortex (left), and temporal pole (left). The seed region of the SA was associated with reduced functional activity with the thalamus (left) and parahippocampal region (left). Further, the seed region of the FPN was associated with increased functional connectivity with the lateral occipital cortex (left) and frontal pole (left), and reduced connectivity with the paracingulate gyrus (bilaterally) and superior frontal gyrus (bilaterally); see **Figures 3** and **eFigure2**.

**Figure 3.**
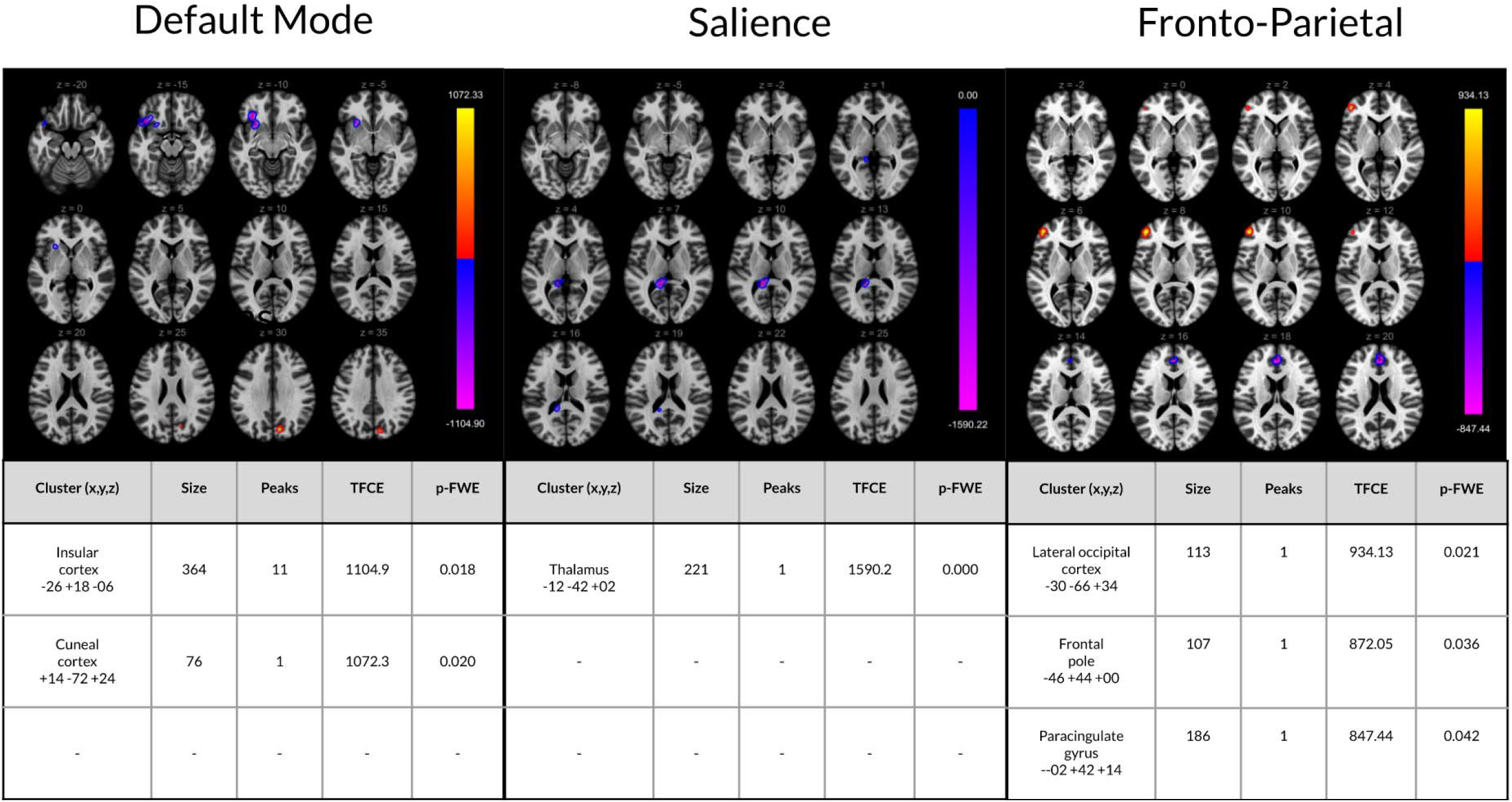
rs-fMRI differences between females with concussion and healthy age-matched females. Clusters (x, y, z) denote standard MNI coordinates at the center of cluster mass, and size represents number of voxels. Only statistics for clusters that survived a p-FWE <0.05 per TFCE are displayed.

In females, groupwise ROI-to-ROI analyses showed that there was increased connectivity between ROIs in the cuneal cortex and cerebellum, as well as between the cingulate gyrus and cerebellar regions. There was also reduced connectivity between the parahippocampal gyrus (posterior division, right) and both the medial pre-frontal cortex and frontal medial cortex in girls with concussion compared to healthy girls (**Figure 4**).

**Figure 4.**
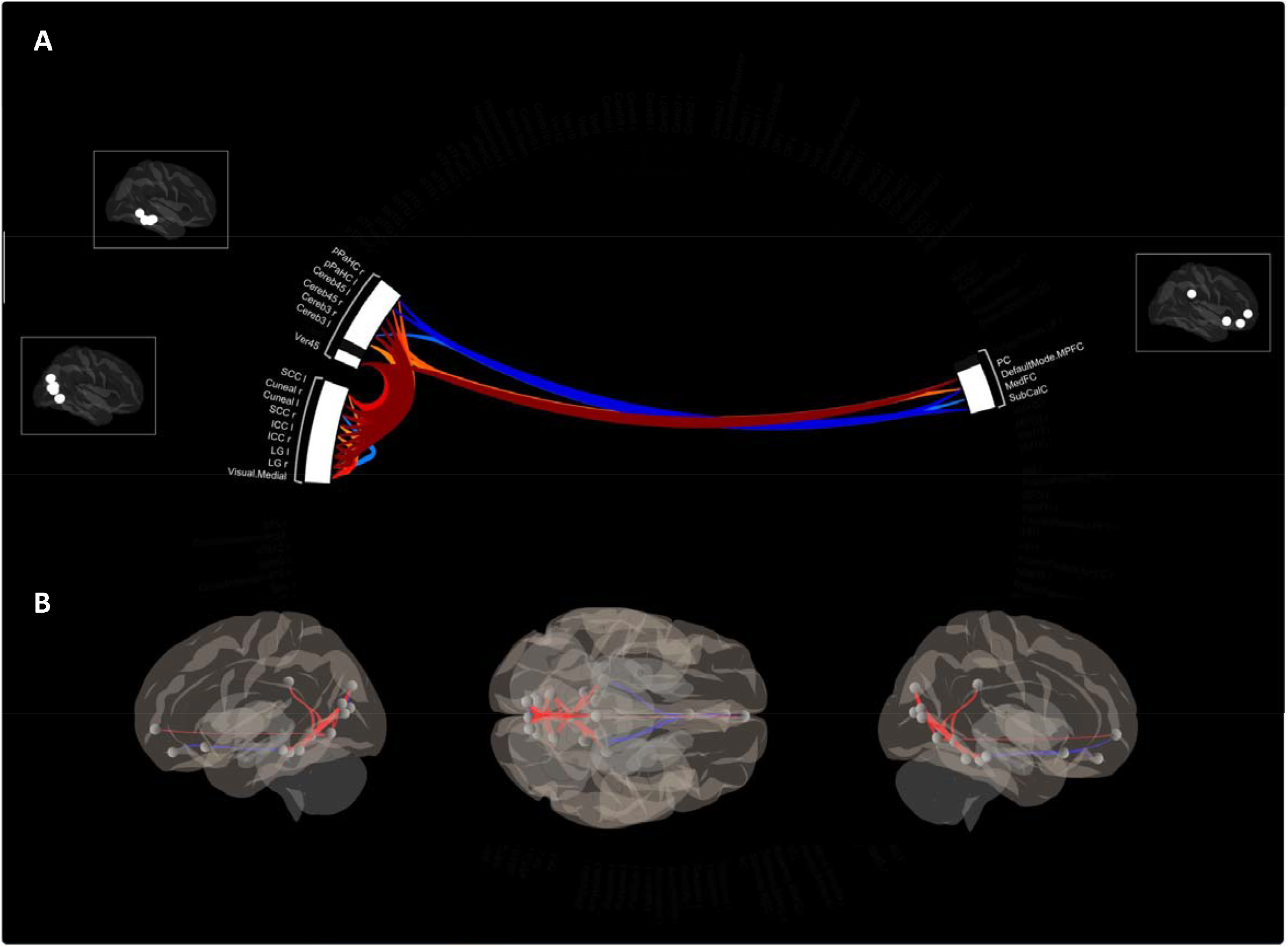
(A) Significantly increased (warm colours) and decreased (cool colours) ROI-to-ROI connectivity in females with concussion in comparison to healthy female controls. (B) Depiction of the ROI-to-ROI changes summarized in panel A on a 3D glass brain to more clearly depict altered patterns of connectivity in an anatomically relevant space.

## DISCUSSION

Our study is the first to report that in pediatric concussion, there are rs-fMRI disturbances observed in females that are not present in the males. To date, the majority of studies on rs-fMRI in pediatric concussion have either not studied females or had a small female representation (<25%) in their samples. Our findings, therefore, provide the first insight into functional disturbances in pediatric concussion by sex.

In pediatric concussion, studies have attempted to link rs-fMRI disturbance to symptoms but have not found a clear relationship^14,18,23^. This may in part be attributable to symptom and rs-fMRI data not being disaggregated by sex in prior studies that have studied both of these measures. Our results show that following concussion, at approximately one-month post-injury, there are alterations in rs-fMRI activity in females (in comparison to their healthy peers) that are not observed in males. Symptom studies align with this, reporting sex-differences with respect to symptoms in pediatric concussion^28,29,31-33^. With a large evidence-base suggesting that symptom presentation differs in pediatric concussion by sex, and with the current study demonstrating sex-based rs-fMRI differences in children with concussion, there is reason to hypothesize that this variable symptom presentation has an underlying functional sex-specific neuropathology. Past studies that did not find a clear relationship between concussion symptoms and functional brain pathology may not have observed such an effect because their analyses were not stratified by sex, which (as shown in the present study) provides insights that mixed-sex analyses do not. Future studies should collect data on symptoms and rs-fMRI and stratify analyses by sex to better understand the relationship between these variables.

Data on the risk of secondary concussion by sex are limited, with the majority of studies to date focusing on predominantly male samples^51^. Our study, however, shows that in females, concussion can impact regions of the brain including the insular cortex, cuneal cortex, and thalamus, which are involved in processing of sensory and/or visual information as well as motor control^52^. These findings may suggest that females, particularly in a sport-related context, may be at elevated risk of secondary injury (owing to potential sensory and/or motor impairments). While data on whether brain function remains impaired at medical clearance to return to activity are mixed^53,54^, our findings (wherein imaging was performed, on average, at the time when clinical recovery from concussion is expected to occur) suggest that functional brain impairments persist in females but not males. This suggests that resolution of functional pathologies in sensory and motor areas of the brain may be sex-dependent, and that return-to-sport guidelines stand to be informed by sex-specific data.

A recent review on sex differences in concussion (pediatric and adult) identified that injury may lead to alterations in the hypothalamic-pituitary-ovarian axis, and subsequent hormonal fluctuations that may be responsible for the more severe symptoms in females^55^. rs-fMRI studies in other endocrinological populations demonstrate that alterations in functional brain activity are associated with abnormal hormonal responses^56-59^. With our study pointing to rs-fMRI disturbances in females with concussion that are not present in males, these functional brain changes may mediate or be related to a variable hormonal response that has an ultimate impact on the female concussion symptomology. Research that directly examines the relationship between brain activity, hormonal fluctuations, and symptomatology is required to build on this possibility.

Large-scale studies (such as the Philadelphia Neurodevelopmental Cohort [PNC], which included nearly 1600 imaging assessments on those aged 8 to 21 years^60,61^) have shown that rs-fMRI patterns vary by sex throughout neurodevelopment. Other studies have also demonstrated sex differences with respect to functional brain activity (and brain morphology, more broadly), and that these differences relate to variable neurodevelopment of networks such as the DMN^62^. In our analyses, we compared concussed males and females to their respective age- and sex-matched control groups, thereby avoiding the potential confounding neurodevelopmental effects that may arise when comparing males to females directly.

## LIMITATIONS AND FUTURE DIRECTIONS

Future studies should include longitudinal assessments to determine if sex-differences in rs-fMRI activity in pediatric concussion vary from acute to the chronic stages of injury. This line of research, combined with existing evidence on pediatric symptom trajectories post-concussion, would help in understanding whether there is a functional neuropathology driving the symptom response longitudinally in children who have delayed recoveries. These studies should also perform rs-fMRI assessments in children who are asymptomatic, given that the broader literature has shown that neurophysiological disturbances can outlast symptoms^63^; understanding whether functional neuropathology outlasts clinical recovery can improve our understanding of the vulnerability of the brain to secondary injuries in a sex-specific manner.

## CONCLUSIONS

This is the first study to report on sex-specific rs-fMRI differences in pediatric concussion. At one-month post-injury, we report on differences in females with concussion (in comparison to their healthy peers) that are not apparent in males. This research further speaks to the need for more sex-specific analyses in concussion research.

## Data Availability

Data are available upon reasonable request.

## FIGURE LEGENDS

**eFigure 1**. For each significant cluster (per TFCE), the effect size associated with that cluster, by group, is depicted below. An associated independent samples t-test (with 52 degrees of freedom) compares the effect sizes at each cluster between groups. Blue and red bars indicate effect sizes for the concussion and control cohorts, respectively.

**eFigure 2**. For each significant cluster (per TFCE), the effect size associated with that cluster by group (healthy females vs. females with concussion) is depicted below. An associated independent samples t-test (with 25 degrees of freedom) compares the effect sizes at each cluster between groups. Blue and red bars indicate effect sizes for the concussion and control cohorts, respectively.

