## Supplementary figures and images for "Sex-specific differences in resting-state functional brain activity in pediatric concussion"

### e-Figure 1

**eFigure 2**

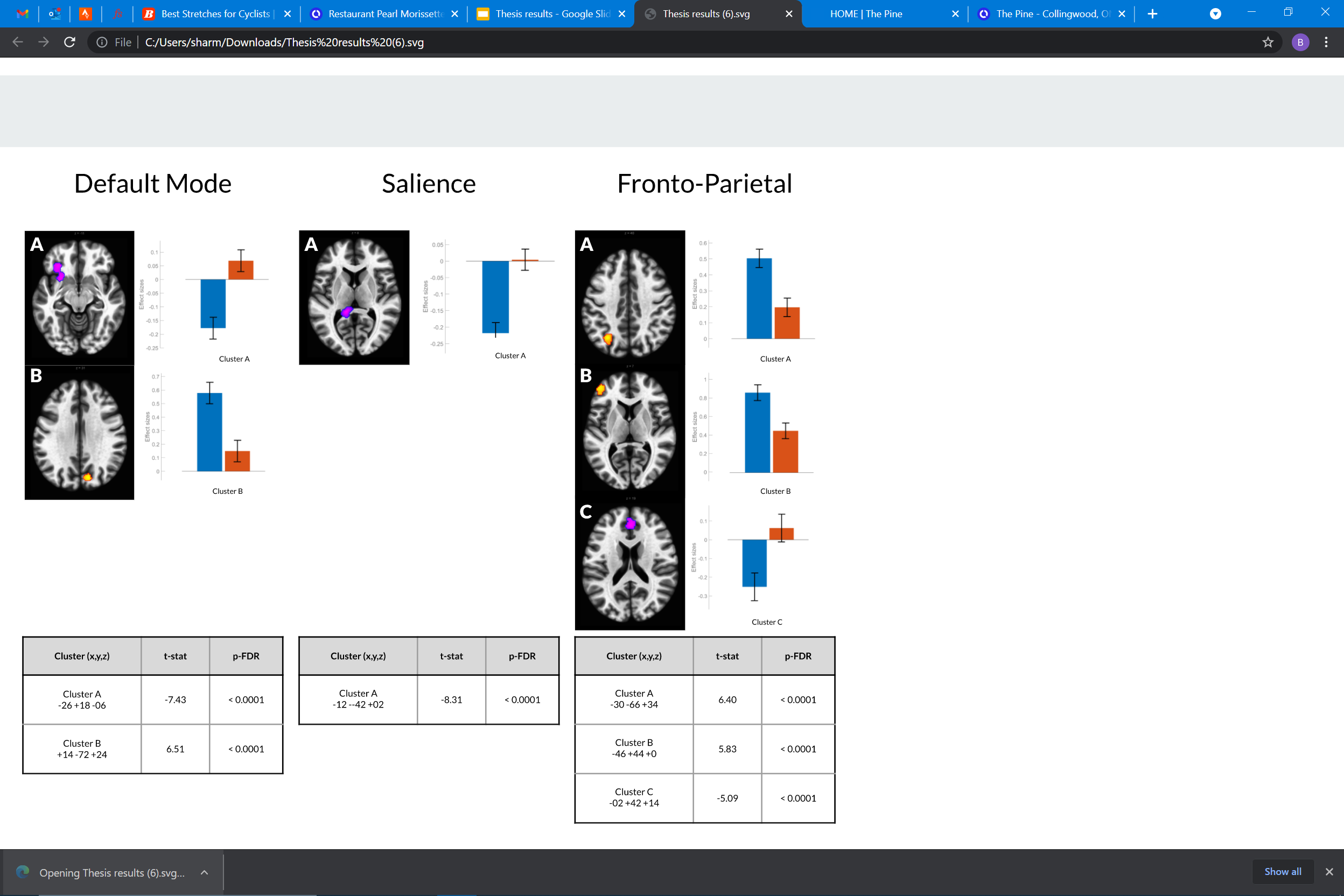

### e-Figure 2

**eFigure 1**

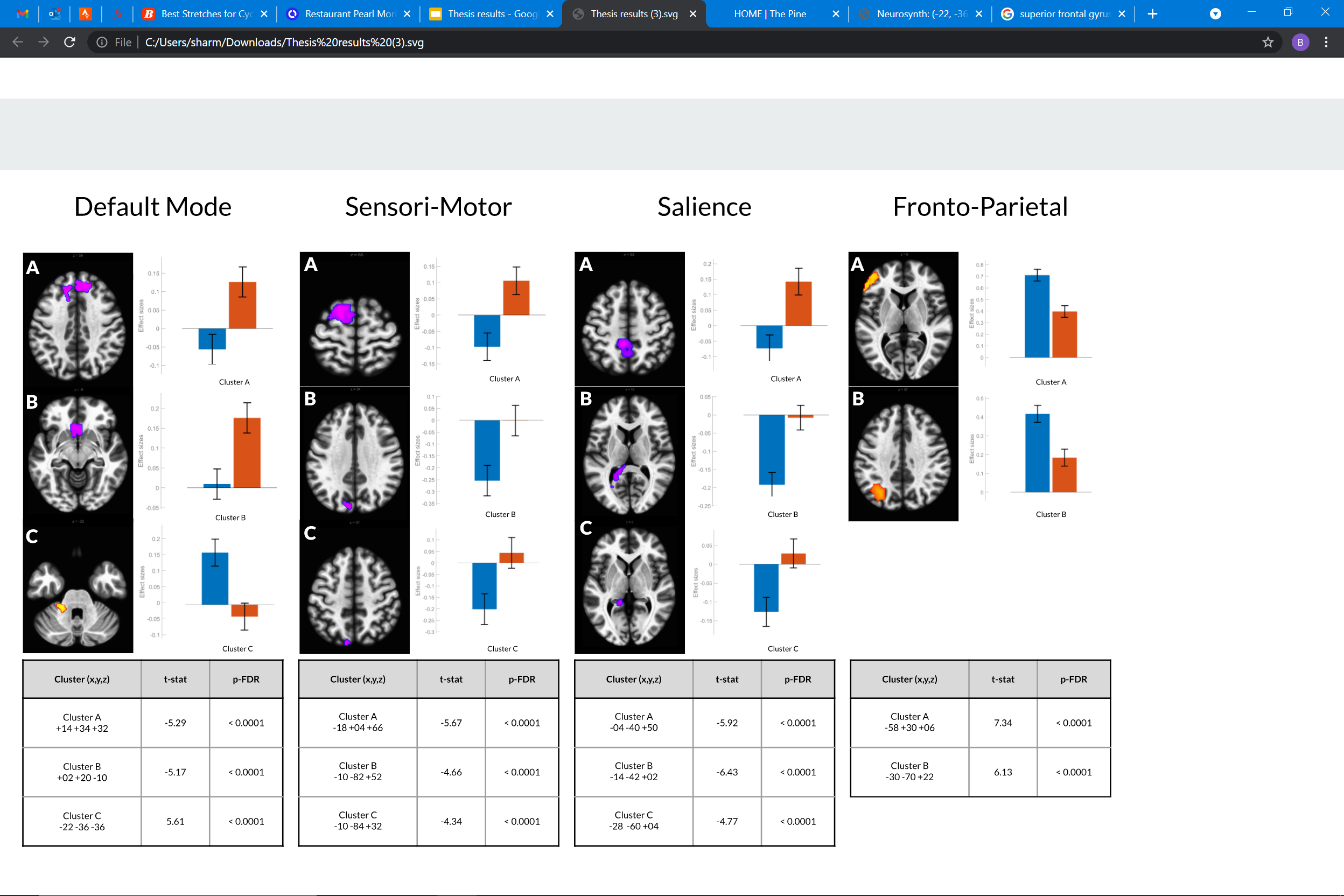
